# Monitoring SARS-CoV-2 in sewage: toward sentinels with analytical accuracy

**DOI:** 10.1101/2021.06.30.21259751

**Authors:** David Calderón-Franco, Laura Orschler, Susanne Lackner, Shelesh Agrawal, David G. Weissbrodt

## Abstract

The severe acute respiratory syndrome coronavirus 2 (SARS-CoV-2) pandemia has been one of the most difficult challenges humankind has recently faced. Wastewater-based epidemiology has emerged as a tool for surveillance and mitigation of potential viral outbreaks, circumventing biases introduced by clinical patient testing. Due to the situation urgency, protocols followed for isolating viral RNA from sewage were not adapted for such sample matrices. In parallel to their implementation for fast collection of data to sustain surveillance and mitigation decisions, molecular protocols need to be harmonized to deliver accurate, reproducible, and comparable analytical outputs. Here we studied analytical variabilities linked to viral RNA isolation methods from sewage. Three different influent wastewater volumes were used to assess the effect of filtered volumes (50, 100 or 500 mL) for capturing viral particles. Three different concentration strategies were tested by electronegative membranes, polyethersulfone membranes, and anion-exchange diethylaminoethyl cellulose columns. To compare the number of viral particles, different RNA isolation methods (column-based *vs*. magnetic beads) were compared. The effect of extra RNA purification steps and different RT-qPCR strategies (one step *vs*. two-step) were also evaluated. Results showed that the combination of 500 mL filtration volume through electronegative membranes and without multiple RNA purification steps (using column-based RNA purification) using two-step RT-qPCR avoided false negatives when basal viral load in sewage are present and yielded more consistent results during the surveillance done during the second-wave in Delft (The Hague area, The Netherlands). By paving the way for standardization of methods for the sampling, concentration and molecular detection of SARS-CoV-2 viruses from sewage, these findings can help water and health surveillance authorities to use and trust results coming from wastewater based epidemiology studies in order to anticipate SARS-CoV-2 outbreaks.

**Graphical Abstract:** 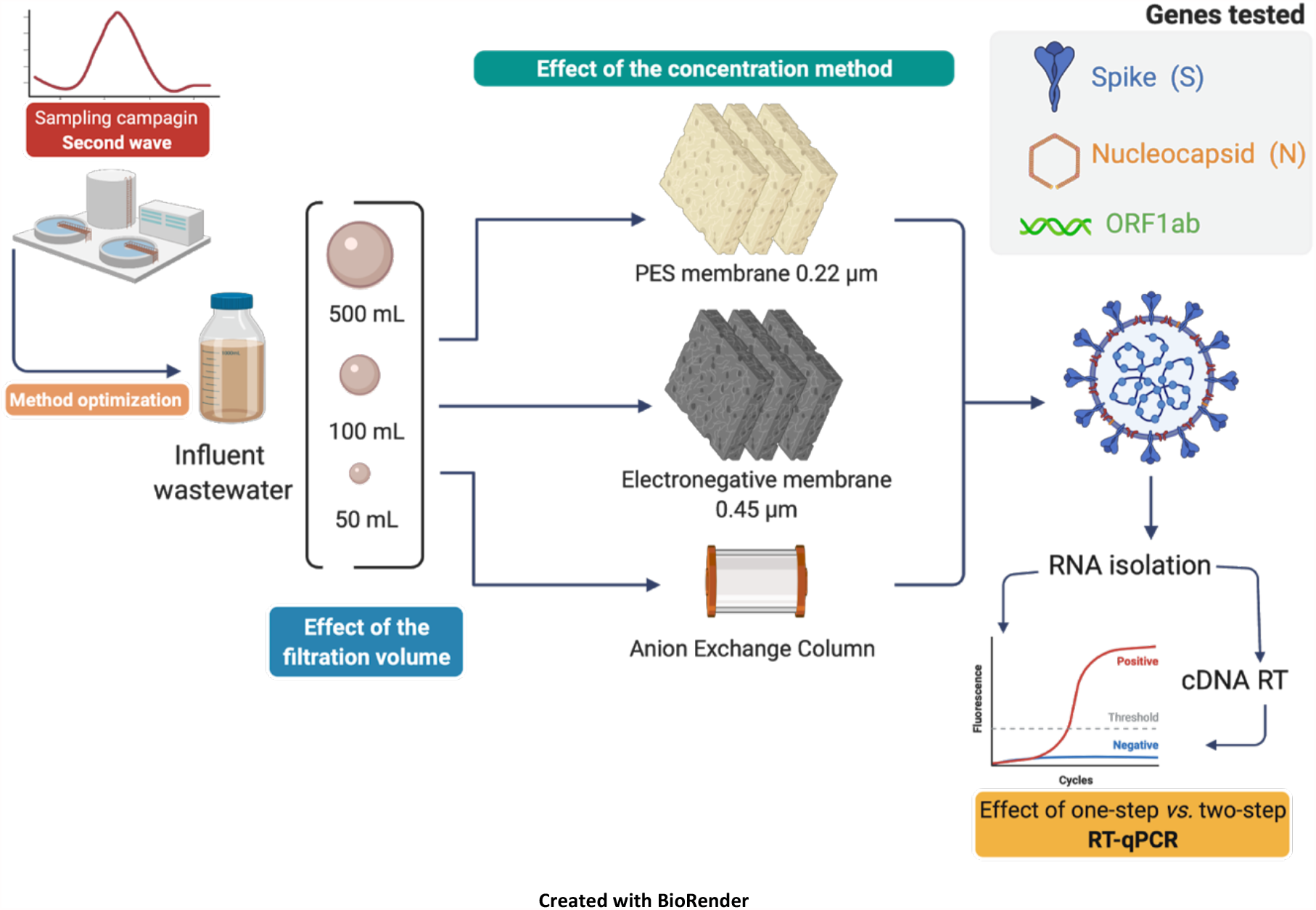

## 1. Introduction

In December 2019, China reported the outbreak of a novel coronavirus named SARS-CoV-2 (genus Betacoronavirus, family Coronaviridae), which has spread around the world fast and lethally. The progression of this SARS-CoV-2 pandemic has been monitored primarily by clinically testing symptomatic individuals for presence of viral RNA. However, as asymptomatic persons seem to account for 40 - 45% of the infections (Ooi and Low, 2020), registering solely symptomatic cases does not allow for prediction of the real cumulative incidence of the viral outbreak. In addition, the number of detected cases highly depends on access to diagnostics as well as the threat of isolation and quarantine dissuading people from getting tested. As SARS-CoV-2 RNA can be found in patients’ stool, samples of rectal swabs, urine and other bodily fluids, detection of the virus via analysing wastewater has gained interest (Agrawal et al., 2021; Foladori et al., 2020; Kitajima et al., 2020; Medema et al., 2020b; Randazzo et al., 2020a; Wu et al., 2020).

Wastewater-based epidemiology can serve as a tool to trace the circulation of a virus variant within a community or municipality. It does not only protect the anonymity of community-members, but it is also unbiased and focused on real-time information, becoming an early-warning system for viral surveillance (Larsen and Wigginton, 2020). It can help make better-informed decisions in public health policies. The urgency of the pandemics pressed the need for a rapid deployment of available analytical methods to track the fate of the virus and its variants. This resulted in a large palette of different protocols for every step. For instance, sampling volumes used vary from a minimum of 2 mL (Rimoldi et al., 2020), to 45 - 60 mL (Hokajärvi et al., 2021; Westhaus et al., 2021), to 100-200 mL (Medema et al., 2020b; Tanhaei et al., 2021), up to 1L (Agrawal et al., 2021); concentration methods vary from precipitation, to centrifugation, ultrafiltration, conventional filtration, filtration by negatively charged membranes, and a combination of these approaches (Kitajima et al., 2020). There is a significant lack of quality controls, variable testing, and methodology improvement that would be necessary to confidently provide analytical accuracy. The global need for comparing virus levels across different communities underlines the need for a standardized workflow (Kitajima et al., 2020). Fortunately, different groups across the world are addressing this issue in order to overcome previous limitations. Protocol improvements have so far been focused in testing different RNA extraction methods (Ambrosi et al., 2021) and on improving the viral concentration steps (Parra Guardado et al., 2020). It has been previously mentioned that one of the variables that should be checked is the filtered volume: low sample volume in combination with low recovery yields could lead to false negative or lower measured SARS-CoV-2 concentration (Alygizakis et al., 2021).

Here, we aimed to establish a method to quantify SARS-CoV-2 from influent wastewater. The effects of filtration volumes of influent wastewater (50, 100 and 500 mL), different concentration methods (polyethersulfone membranes, electronegative membranes and anion-exchange chromatography), different RNA isolation methods (column based *vs*. magnetic beads), and different RT-qPCR strategies (one-step *vs*. two-step) were assessed. A methodology designed from the selected best parameter testing outputs was used to survey viral gene copies across the second wave of the pandemic in the Delfland water catchment area (The Hague, The Netherlands) and compare it to publicly available clinical data. We deliver an assessment of method variability and biases introduced when basal viral loads are present and establish a methodology implementable in wastewater-based epidemiology. This will support health and water authorities to anticipate, monitor, and control future outbreaks.

## 2. Material and Methods

A summary of the methodology can be found in **Figure 1**. In brief, municipal wastewater was collected to test the effect of 3 filtration volumes (50, 100, 500 mL) and 3 virus concentration methods (polyethersulfone membrane, electronegative membrane, anion exchange column) on molecular analyses of 3 genes (spike S, nucleocapsid N, ORF1ab) of SARS-CoV-2 by one-step and two-step reverse transcription (RT) and quantitative PCR (qPCR) after isolation of viral RNA using single (magnetic beads, columns) and multiple purification methods. The optimal method parameters were then applied to monitor a time series over the second wave of the epidemics.

**Figure 1.**
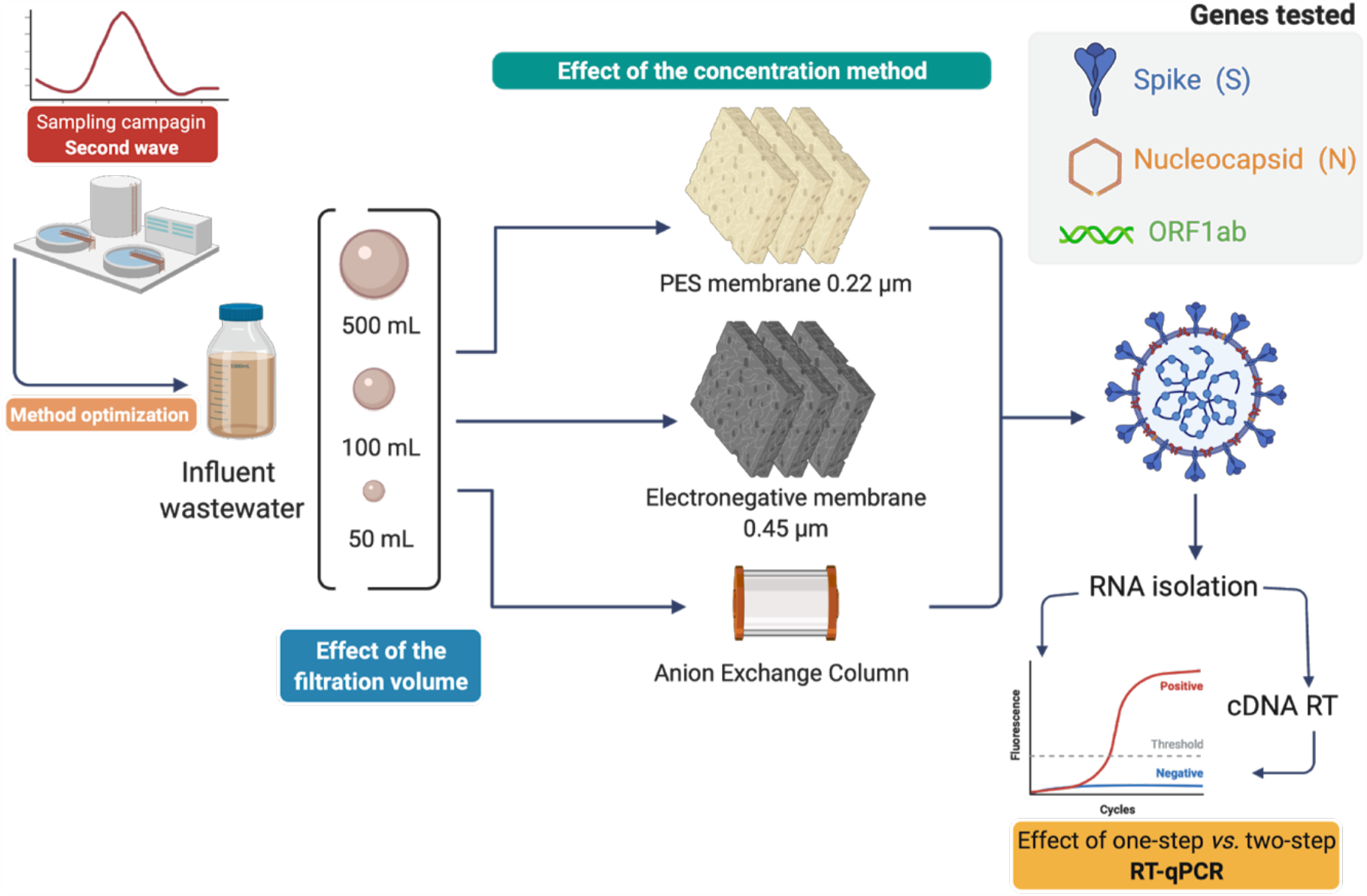
Schematic representation of the experimental design followed in this study. Viral genes (S, N and ORF1ab) were quantified by RT-qPCR (one-step vs two-step RT-qPCR) after testing the effect of the filtration volume and different concentration methods. Created with BioRender.

### 2.1. Sampling of influent water from WWTP

Influent wastewater was collected before primary treatment from WWTP Harnaschpolder (Delft, The Netherlands). Volumes of 5 L from 24-h flow-proportional composite samples were collected per time point. Samples were obtained every 3 weeks over 6 months from July 7 to December 1, 2020 covering the end of the first wave and the whole second wave of COVID-19 in The Netherlands (**Figure 2**). The 24-composite sample from July 7 2020 was used for method development. All samples were processed in a timeframe of less than 1 day after collection.

**Figure 2.**
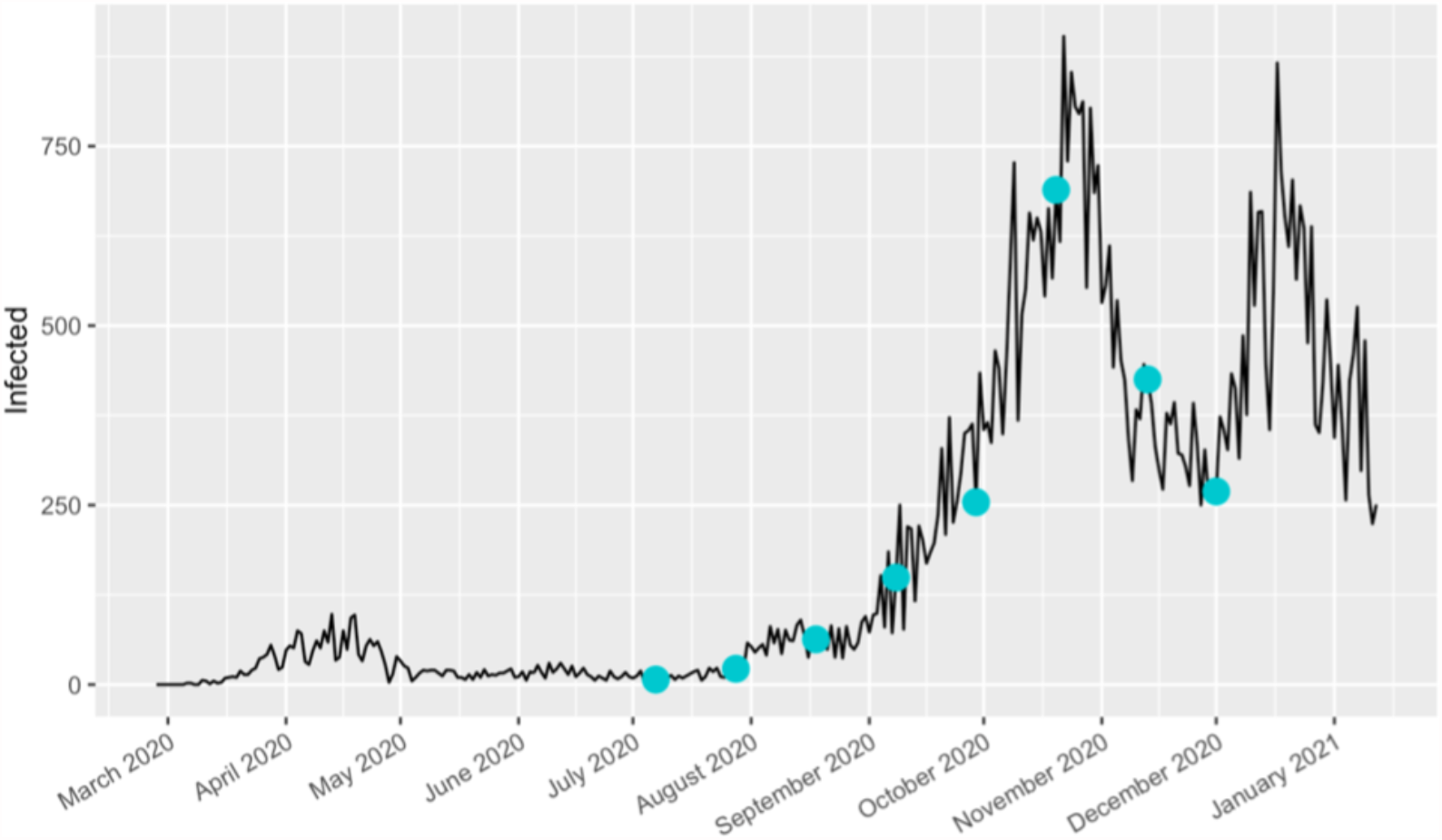
Graphical representation of the number of positive cases of COVID-19 cases in The Hague area (black line) and the dates where samples were taken (blue dots) (Data source = Coronadashboard-RIVM https://coronadashboard.rijksoverheid.nl).

### 2.2. Deactivation of SARS-CoV-2

Influent wastewater samples were heated at 65°C for 30 minutes to deactivate and handle the sewage sample under biosafety conditions (Batéjat et al., 2021; Rabenau et al., 2005).

### 2.3. Concentration of SARS-CoV-2 from wastewater

The effect of used influent wastewater volume and the concentration method was assessed for viral genes quantification. A 24-h composite sample (July 7 2020) was used, representing the beginning of the second wave **(Figure 2)**. The goal was to identify the best concentration method when basal viral levels were present in influent wastewater. A volume of 200 µL (50 ng µL^-1^) of an F-specific RNA MS2 phage (Thermo Fisher Scientific, Germany) was spiked as internal standard in the influent wastewater solution before the concentration methods. The group of F specific RNA phages (f2 and MS2 are the best known members of this group) are considered the most attractive subgroup of phages to serve as indicator organisms because their physical structure closely resembles that of enteroviruses in wastewater (Havelaar et al., 1985).

#### 2.3.1. Effect of the volume of influent wastewater used

Volumes of 50, 100 and 500 mL of influent wastewater were tested. All the analyses were performed in three replicates.

#### 2.3.2. Effect of methods used to concentrate virus particles

These influent wastewater volumes were filtered (membranes) or loaded (chromatographic column) in three technical replicates with three different methodologies: (i) filtration on polyethersulfone (PES) membrane of 0.22 µm pore size (Pall Corporation, USA); (ii) retention on a 1-mL diethylaminoethyl cellulose (DEAE) anion-exchange chromatography (BIA separations, Slovenia) which is used for phages separations (Kattur Venkatachalam et al., 2014; Kramberger et al., 2010) and for isolations of free-floating extracellular DNA from water matrices (Calderón-Franco et al., 2021); and (iii) attraction on electronegative membranes of 0.45 µm pore size (Merck Millipore, The Netherlands). Prior to loading on the DEAE chromatography column, the influent wastewater was filtered through the 0.22 µm PES to remove biosolids and particles, which is necessary to protect the DEAE column. Loading and elution in chromatography is more time consuming than direct filtration. However, chromatography is more selective and leads to higher grades of purity of concentrated analytes. Thus, we also considered the DEAE method as a way to identify the recovery of viral RNA when compared to the filtration methods.

### 2.4. Effect of isolations and purifications of the RNA of SARS-CoV-2

Extractions of raw viral RNA from membrane filters and DEAE column eluents were performed by using two different commercial kits for RNA extraction. First, we used the MagMax CORE Nucleic Acid Purification kit (Thermo Fisher Scientific, The Netherlands), that employ magnetic beads for nucleic acid purification, together with the MagMax CORE Mechanical Lysis Module (Thermo Fisher Scientific, The Netherlands) by following manufacturer’s recommendations. Second, we used the Fast RNA Blue kit (MP Biomedicals), which is based on column-based purification (solid phase extraction), according to the manufacturer’s protocol. In addition, the effect of multiple RNA purification steps before quantification of genes by RT-qPCR was assessed by involving the GeneJET NGS Cleanup Kit (Thermo Scientific, USA) according to manufacturer’s instructions obtaining the purified RNA sample.

The quality and quantity of raw and purified RNA extracts were measured with a NanoDrop Spectrophotometer (ND-1000,USA).

### 2.5. Effect of one-step *vs*. two-step RT-qPCR on detection of basal RNA concentrations

On the lowest basal viral load day from our sampling campaign (7th July 2020), we compared one-step and two-step RT-qPCRs in order to assess the effect of quantification of viral RNA genes when the incidence of cases is low. This was assessed to avoid potential false negatives. One-step RT-qPCR was performed using the isolated RNA as a template (raw RNA), which included specific primers for the target genes. Two-step RT-qPCR was performed by synthesizing complementary DNA (cDNA) from the pool of raw RNA isolated per sample using the SuperScript VILO cDNA synthesis kit (Thermo Fisher Scientific, The Netherlands) following manufacturer’s recommendations, included random primers, prior to using the cDNA as a template for qPCR. The concentration of the synthesized cDNA was measured with Qubit® dsDNA assays (Thermo Fisher Scientific, USA).

### 2.6. Detection and quantification of SARS-CoV-2 RNA by RT-qPCR

We quantified the concentration of SARS-CoV-2 RNA in wastewater samples by measuring marker genes using the TaqPath COVID-19 RT-PCR Kit (Thermo Fisher Scientific, Germany) with a QuantStudio 3 Thermal Cycler, according to a previous study (Agrawal et al., 2021). The TaqPath COVID-19 RT-PCR Kit includes primer pairs targeting genes that code for the structural transmembrane spike (S) glycoprotein and nucleocapsid (N) protein, and for the non-structural open reading frame of the replicase complex (ORF1ab) and that were used in a multiplex assay (**Table S1**). Details about the kit are provided in the Supplementary Information. Each qPCR run was performed in technical triplicates in reaction volumes of 50 µL, with 12.5 µL TaqPath 1-Step Multiplex Master Mix (4X), 2.5 µL COVID-19 Real Time PCR Assay Multiplex Diagnostic Solution, and 25 µL nuclease free water. To the reaction mix, 10 µL of purified and extracted viral RNA were added. Thermal profiles are provided in Supplementary Information (**Table S2**). Reactions were considered positive if the cycle threshold (CT) was below 40 cycles, otherwise negative (*i*.*e*., no detection of the SARS-CoV-2 RNA in the sample). The lower limit of detection was 10 gene copies per RT-qPCR reaction.

### 2.7. Statistics

Statistical analyses were performed on all molecular datasets with R 3.5.1 (R Foundation for Statistical Computing., 2018) and RStudio (https://www.rstudio.com/). The RT-qPCR abundance data were analyzed in R using ggplot2 (v0.9.3.1).

## 3. Results and Discussion

The effect of different initial influent wastewater volumes (50, 100 and 500 mL), different viral concentration methods, different RNA purification methods, and different one-step/two-step RT-qPCR methods were assessed on a basal viral level timepoint (July 7 2020), corresponding to the beginning of the second wave (**Figure 2**) and to the lowest day in terms of cumulative incidence of COVID-19 cases. This timepoint served as reference for method standardization as basal viral load levels from influent wastewater samples. The selected methodology was further used for surveilling SARS-CoV-2 gene copies over the second wave (July to December 2020).

### 3.1. Electronegative membranes and larger volumes were optimal to quantify SARS-CoV-2 from sewage samples

Among the three different methods tested to concentrate virus particles (electronegative membranes, PES membranes, DEAE column), concentration by electronegative membranes of 0.44 µm pore size was the only one displaying positive results in terms of detection of S, N, ORF1ab genes and MS2 phage control **(Figure 3)**.

**Figure 3.**
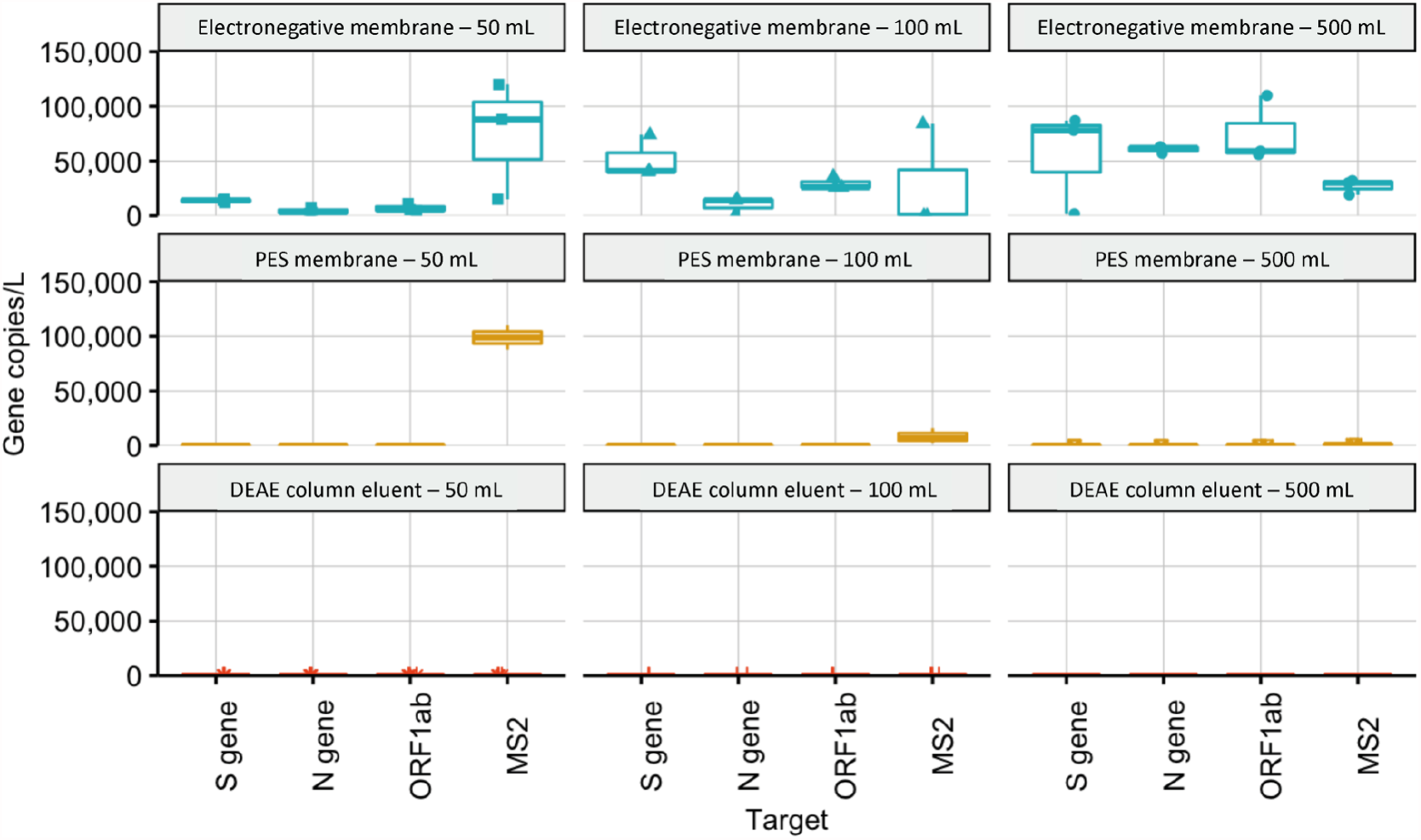
Concentration of SARS-CoV-2 RNA in influent wastewater as determined by the N, ORF1ab,S and MS2 control gene assays in gene copies per liter, using different isolation methods (electronegative membranes filtration, polyethersulfone membrane (PES) filtration and diethylaminoethyl cellulose (DEAE) column) and different initial volumes (50, 100 and 500 mL).

When PES membranes were used, SARS-CoV-2 was not detected from the cake, suggesting that there was no retention to the membrane. However, internal control MS2 phage, which was added prior to filtration, was detected in PES membrane. This difference may result from structural viral conformations such as particle sizes of 100 nm (SARS-CoV-2) *vs*. 20 nm (MS2) or if virus is enveloped (SARS-CoV-2) *vs*. non-enveloped (MS2), resulting in different bindings to the membrane materials (Rockey et al., 2020).

The permeate of PES membranes was subsequently loaded on a DEAE chromatographic column to verify if the SARS-CoV-2 virus particles could be adsorbed and isolated similar to phages (Kattur Venkatachalam et al., 2014; Kramberger et al., 2010) and free-floating exDNA (Calderón-Franco et al., 2021). However, the DEAE method did not result in concentrating SARS-CoV-2, suggesting that neither the PES membrane, nor the DEAE column is suitable for SARS-CoV-2 concentration from wastewater.

SARS-CoV-2, among other viruses, has an isoelectric point (pI = ½ (pKa_1_ + pKa_2_) below 7. The pH of the influent wastewater pH ranged between 6.3 - 6.8. At pH values below the isoelectric point, the charge of SARS-CoV-2 and MS2 phage could be positive, thus explaining why they did not bind to the DEAE (positive net charge) column and did bind to the electronegative membranes (Joonaki et al., 2020).

When expressing the RT-qPCR results in concentrations of viral gene copies detected per L, filtering 500 mL influent water through electronegative membranes gave, in average, higher values of gene copies per L (7.2·10^4^ ± 1.7·10^4^ viral gene copies L^-1^) when compared to gene copies obtained from 50 mL (7.4·10^3^ ± 4.1·10^3^ viral gene copies L^-1^) or 100 mL (3.0·10^4^ ± 1.9·10^4^ viral gene copies L^-1^). In **Figure 3**, individual gene results were also more consistent when 500 mL were filtered (S gene: 5.6·10^4^ ± 2.8·10^4^ viral gene copies L^-1^; N gene: 6.0·10^4^ ± 2.6·10^3^ viral gene copies L^-1^; ORF1ab gene: 7.5·10^4^ ± 2.4·10^4^ viral gene copies L^-1^) when compared to filtered 50 mL (S gene: 1.3·10^4^ ± 1.0·10^4^ viral gene copies L^-1^; N gene: 4.2·10^3^ ± 2.3·10^3^ viral gene copies L^-1^; ORF1ab gene: 6.9·10^3^ ± 2.9·10^3^ viral gene copies L^-1^) and 100 mL (S gene: 5.2·10^4^ ± 1.6·10^4^ viral gene copies L^-1^; N gene: 9.7·10^3^ ± 6.7·10^3^ viral gene copies L^-1^; ORF1ab gene: 2.9·10^4^ ± 4.7·10^3^ viral gene copies L^-1^).

Here we demonstrated that filtration of larger volumes (*i*.*e*., 500 mL) in combination of electronegative membranes allowed, in the lowest day of COVID-19 incidence of the second wave, to adsorb and concentrate enough SARS-CoV-2 for highest and most consistent (uniform results among gene tested) detection and quantification by two-step RT-qPCR.

### 3.2. Avoiding multiple RNA purification steps and performing two-step RT-qPCR resulted in higher viral quantification

One of the major analytical concerns related to the wastewater-based epidemiology of SARS-CoV-2 is the quality and purity of the isolated RNA, especially when low viral loads are present. This can potentially compromise the accuracy of the results obtained through false negatives. The effect of multiple RNA purification steps and one-step/two-step RT-qPCR strategies were assessed. The best concentration method obtained during the first phase of this study (*i*.*e*., 500 mL influent wastewater filtered on electronegative membranes) was utilized.

In **Figure 4**, it can be observed that for the three genes tested (N, ORF1ab and S), the two-step RT-qPCR performed from raw and purified RNA displayed, on average, higher values (1.6·10^5^ ± 1.0·10^4^ viral gene copies 500 mL^-1^) than one-step RT-qPCR (no positive results from tested genes). No detection was obtained from one-step RT-qPCR both from raw RNA and purified RNA with the exception of the control MS2 sample, whereas one-step RT-qPCR from purified RNA resulted in the detection of 2.9·10^7^ ± 4.2·10^6^ MS2 gene copies 500 mL^-1^. Raw RNA obtained with magnetic beads contained high concentration of environmental inhibitors that made the binding between RNA and beads difficult during the processing of large volumes of influent wastewater, hampering the qPCR analysis (Graham et al., 2021; Kitajima et al., 2020; Schrader et al., 2012). This can affect the sensitivity of the assay and result in false-negative results (Ahmed et al., 2020).

**Figure 4.**
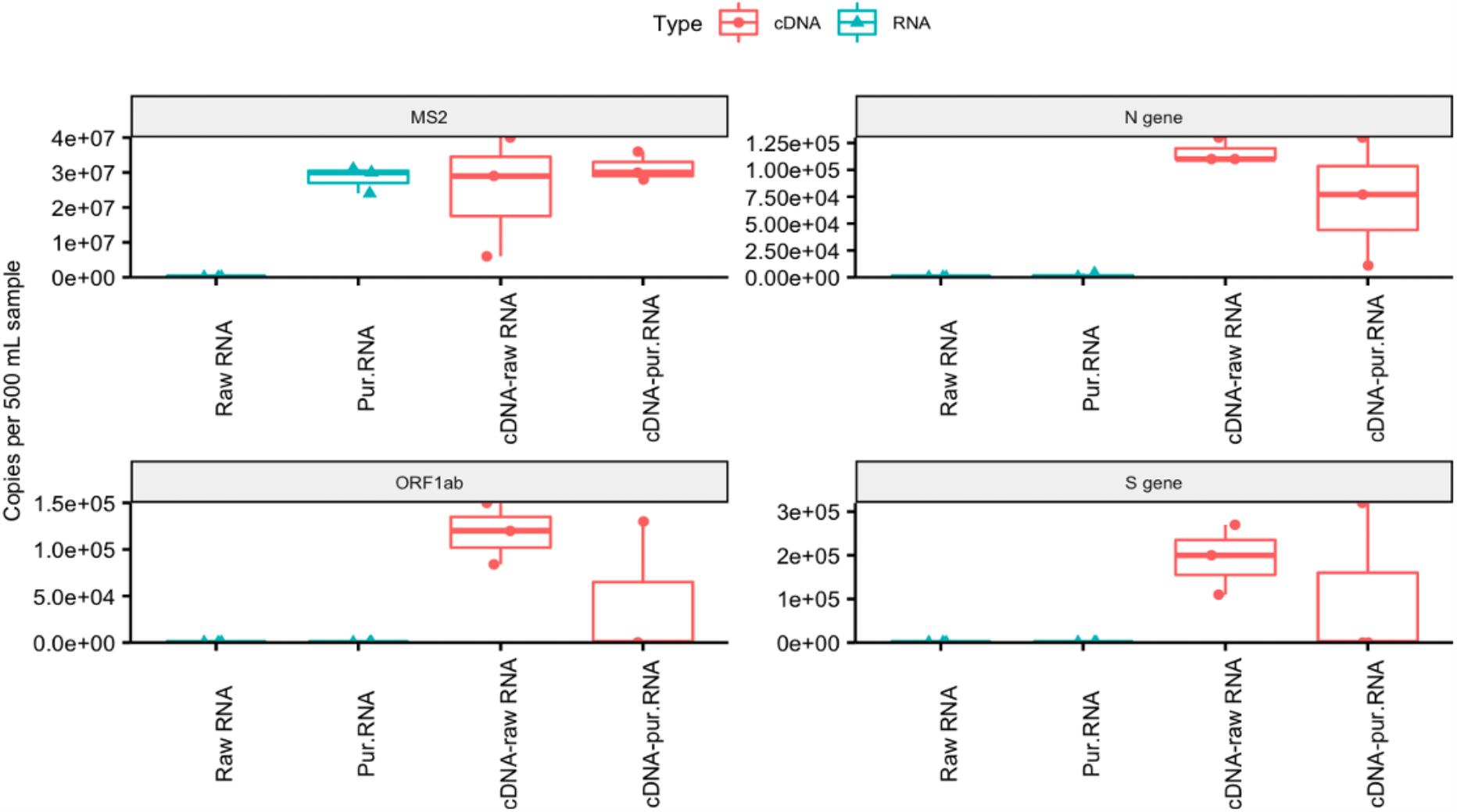
Concentration of SARS-CoV-2 RNA in influent wastewater as determined by the N, ORF1ab, S and MS2 control gene assays in gene copies per 500 mL, using one-step (raw RNA and purified RNA) and two-step (cDNA-raw RNA and cDNA-purified RNA) RT-qPCR.

The three genes were consistently detected by two-step RT-qPCRfrom both raw RNA and purified RNA. However, RNA purification induced higher variability on the average of all the tested genes (1.7·10^5^ ± 1.1·10^5^ gene copies in 500 mL, 65% relative standard deviation (RSD)) when compared to the lower standard deviation obtained with two-step RT-qPCR (1.5·10^5^ ± 3.7·10^4^ gene copies in 500 mL, 25% RSD). When individual genes were tested, the N gene exhibited the highest variability (2-fold) between raw (1.2·10^5^ ± 8.0·10^4^ gene copies in 500 mL) and purified (7.3·10^4^ ± 6.0·10^4^ gene copies in 500 mL) RNA. Thus, multiple RNA purification stages before RT-qPCR are not recommended: RNA from viral particles were lost during the purification process even if detected with two-step RT-qPCR. The same trend was observed during the time course of the 6-months sampling campaign, where two-step RT-qPCR measurements displayed higher and more consistent results (*i*.*e*., uniform values between the three tested genes) (**Figure S1**).

Our observation about how relevant it is to consider one-step *vs*. two-step RT-qPCR when viral load in wastewater samples is very low is in consensus with a previous study (Chik et al., 2021). This was of high importance as it was the day, from the sampling campaign, with the lowest viral load. Chik et al. (2021) also emphasize that, when the viral load is highly concentrated, the effect of one-step *vs*. two-step RT-qPCR is less relevant. However, assessing ways of quantifying RNA while avoiding false negatives are needed to get accurate values to sustain decisions to mitigate outbreaks. Chik et al. (2021) have performed collaborative inter-laboratory experiments where they spiked low and high levels of surrogate SARS-CoV-2 virus. They have observed less variation between laboratories when high-spike conditions were compared to the low-spike conditions. The Ct value Chik et al. (2021) obtained from analysis of the surrogates in the low-spike samples was not in the linear range of PCR amplification and approached the sensitivity limit. Therefore, it is recommended to: (1) avoid multiple RNA purification steps for reducing loss of the RNA; (2) and run a two-step RT-qPCR analysis, which allow enhancing the resolution. This is because cDNA is firstly synthesized from total viral RNA using random primers followed by specific primers for qPCR analysis, being able to detect low-levels of SARS-CoV-2 RNA copies in sewage.

### 3.3. Wastewater-based epidemiology as a tool to anticipate viral outbreaks

The range of commercial options to isolate RNA and quantify SARS-CoV-2 viral particles is diverse and complex. Two different RNA isolation methods were chosen to survey the number of viral particles over the sampling campaign **(Figure 5)**: one column-based microbial RNA extraction kit (Fast RNA) and one magnetic beads-based kit (MagMax Core kit).

**Figure 5.**
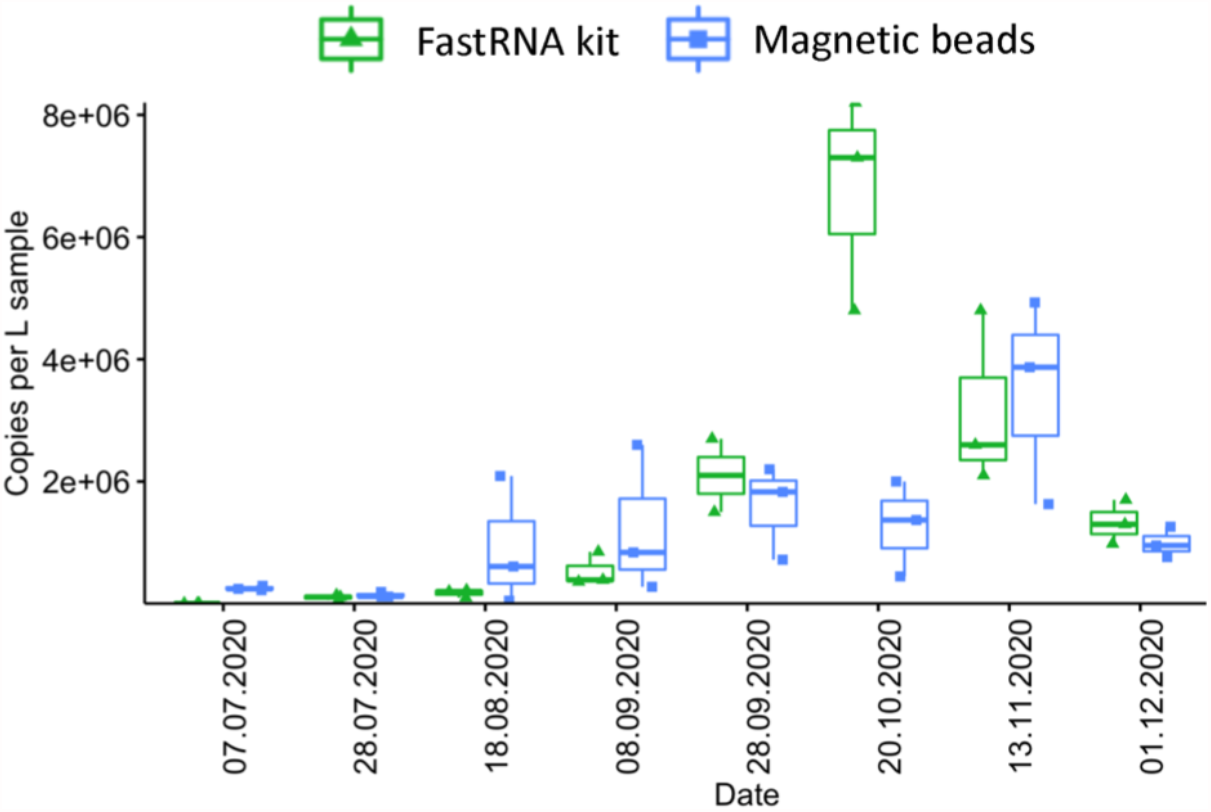
Concentration of SARS-CoV-2 RNA in influent wastewater as determined by the average of N, ORF1ab and S gene assays in gene copies per L using two RNA extraction methods: FastRNA and MagMax (Magnetic beads) over the whole sampling campaign (July 2020 until December 2020) covering the second wave in The Netherlands.

Comparative analysis between RNA extraction methods are displayed in **Figure 5**. Both methods showed similar results in terms of gene copies per liter (no significant difference; p>0.05) during the whole sampling campaign with the exception of October 20, 2020 (p<0.05), which happened to be the peak of the second wave. The column-based method resulted in less variation between replicates when compared to the magnetic beads method. Our results can be compared with the results from a previous measuring campaign performed in The Hague (Medema et al., 2020a), where results were reported as concentrations of N gene copies per mL on the first wave of the pandemics from February 2020 until August 2020. The results of July and August were in the range of 10^4^-10^5^ viral N gene copies L^-1^. The concentrations obtained with our optimized methodology are 10-fold higher (10^5^-10^6^ gene copies L^-1^) taking into account the average of these three genes (N, S and ORF1ab) than those obtained by Medema et al. (2020a). It allowed a more sensitive quantification of viral gene copies at the beginning of the pandemic second-wave.

Early detection of SARS-CoV-2 and other future possible viral agents from wastewater requires accurate and adapted-to-matrix methodologies. Thus, being able to quantify low viral levels with non-clinical samples is an important factor to consider in wastewater-based epidemiology, as early communication to the competent authorities is necessary to have enough reaction time to prevent viral outbreaks in urban settlements.

The concentrations measured matched with the COVID19 incidences reported by the National Institute for Public Health and Environment (RIVM, The Netherlands) and seemed to slightly anticipate by 1-2 weeks the start of the second wave. The second peak started to increase in the beginning of September 2020 while significant viral copies could already be quantified by August 18, 2020 (1.73·10^6^ viral gene copies L^-1^). This is in accordance with Larsen and Wigginton (2020), who have claimed that wastewater-based epidemiology remediates clinical biases that result from low access to isolation diagnostics and quarantine threat dissuading citizens from getting tested. It also helps understand the locations and dynamics of community transmissions, becoming a more cost-effective and less invasive method. Randazzo et al. (2020b) have highlighted the importance of wastewater-based epidemiology as an early indicator, revealing that members of the community were shedding SARS-CoV-2 RNA particles in their stool even before the first cases had been reported by the authorities in Murcia (Spain).

Overall, a global effort is necessary to generate, harmonize, and use standardized analytical protocols for an accurate, reproducible, and comparable detection and quantification of SARS-CoV-2 (Bivins et al., 2020). On a wastewater-based epidemiology basis, crucial considerations are needed from sampling to preparation and measurements in order to define (i) the sampling points, (ii) the sample size and frequency, (iii) the method to efficiently concentrate viral particles, (iv) the quantification methods used, (v) the primer set(s), (vi) the controls, and (vii) the normalization methods. Some initiatives on protocol standardization have been taken such as the NORMAN-SCORE joint initiative to facilitate data comparison between “SARS-CoV-2 in sewages” studies (Lundy et al., 2021). Knowing the inherent biases associated with qPCR analysis of wastewater samples due to the variability in the matrix of the samples (Agrawal et al., 2021; Chik et al., 2021) and the urgency of time, we believe that it is currently important to focus on the robustness and reproducibility of the qPCR method used in each respective lab, especially during low incidence values. Analytical robustness should help translate risk in the form of viral concentrations per volume filtered to react fast enough to minimize undesirable infective and spreading consequences.

## 4. Conclusions

Molecular sentinels require analytical accuracy for inter-comparison and information delivery. From this work, we conclude that:

1. The higher the influent wastewater sample volume was (500 mL instead of 100 or 50 mL), the more uniform the concentrations of the three viral genes (N, S, ORF1ab) were, especially when low incidence were reported
2. Employing a two-step RT-qPCR method would help to enhance the resolution for samples with low viral loads.
3. Electronegative membranes were optimal and cheap for the concentration of SARS-CoV-2 from pre-settled influent wastewater.
4. Multiple RNA purification steps are not recommended, due to plausible loss of SARS-CoV-2 RNA and thereby, affecting qPCR analysis outcomes. While comparing the column-based vs. magnetic beads method for the RNA purification, we found that the column-based purification method displayed less variability amongst the biological triplicates throughout the sampling campaign than magnetic beads.

## Data Availability

All data necessary can be found in supplementary material

## Conflict of interests statement

The authors declare no conflict of interest.

## Authors’ contributions

DCF, SA and DGW designed the study. DCF, SA and LO performed the experimental investigations. DCF wrote the manuscript with direct contribution, edits, and critical feedback by all authors.

## Acknowledgements

We are grateful to Mariska Ronteltap from the Delfland Water Authority and Abdelmaoula Idrissi from Delfluent Services and WWTP Harnashpolder, The Netherlands, for helping us arrange this sampling campaign in such difficult moments, as well as Gertjan Medema from KWR and TU Delft for the comparison of results with the measurement campaign in The Hague. This work is part of the research project “Transmission of Antimicrobial Resistance Genes and Engineered DNA from Transgenic Biosystems in Nature” (Targetbio) funded by the program Biotechnology & Safety (grant no. 15812) of the Applied and Engineering Sciences Division of the Dutch Research Council (NWO).

## Notes

### Competing Interest Statement

The authors have declared no competing interest.

### Author Declarations

We confirm that all relevant ethical guidelines have been followed. All necessary patient/participant consent has been obtained and the appropriate institutional forms have been archived.

## References

Agrawal, S., Orschler, L., Lackner, S., 2021. Long-term monitoring of SARS-CoV-2 RNA in wastewater of the Frankfurt metropolitan area in Southern Germany. Sci. Rep. 11, 5372. https://doi.org/10.1038/s41598-021-84914-2

Ahmed, W., Bivins, A., Bertsch, P.M., Bibby, K., Choi, P.M., Farkas, K., Gyawali, P., Hamilton, K.A., Haramoto, E., Kitajima, M., Simpson, S.L., Tandukar, S., Thomas, K. V., Mueller, J.F., 2020. Surveillance of SARS-CoV-2 RNA in wastewater: Methods optimization and quality control are crucial for generating reliable public health information. Curr. Opin. Environ. Sci. Heal. 17, 82–93. https://doi.org/10.1016/j.coesh.2020.09.003

Alygizakis, N., Markou, A.N., Rousis, N.I., Galani, A., Avgeris, M., Adamopoulos, P.G., Scorilas, A., Lianidou, E.S., Paraskevis, D., Tsiodras, S., Tsakris, A., Dimopoulos, M.A., Thomaidis, N.S., 2021. Analytical methodologies for the detection of SARS-CoV-2 in wastewater: Protocols and future perspectives. TrAC - Trends Anal. Chem. 134, 116125. https://doi.org/10.1016/j.trac.2020.116125

Ambrosi, C., Prezioso, C., Checconi, P., Scribano, D., Sarshar, M., Capannari, M., Tomino, C., Fini, M., Garaci, E., Palamara, A.T., De Chiara, G., Limongi, D., 2021. SARS-CoV-2: Comparative analysis of different RNA extraction methods. J. Virol. Methods. https://doi.org/10.1016/j.jviromet.2020.114008

Batéjat, C., Grassin, Q., Manuguerra, J.-C., Leclercq, I., 2021. Heat inactivation of the severe acute respiratory syndrome coronavirus 2. J. Biosaf. Biosecurity 3, 1–3. https://doi.org/10.1016/j.jobb.2020.12.001

Bivins, A., North, D., Ahmad, A., Ahmed, W., Alm, E., Been, F., Bhattacharya, P., Bijlsma, L., Boehm, A.B., Brown, J., Buttiglieri, G., Calabro, V., Carducci, A., Castiglioni, S., Cetecioglu Gurol, Z., Chakraborty, S., Costa, F., Curcio, S., De Los Reyes, F.L., Delgado Vela, J., Farkas, K., Fernandez-Casi, X., Gerba, C., Gerrity, D., Girones, R., Gonzalez, R., Haramoto, E., Harris, A., Holden, P.A., Islam, M.T., Jones, D.L., Kasprzyk-Hordern, B., Kitajima, M., Kotlarz, N., Kumar, M., Kuroda, K., La Rosa, G., Malpei, F., Mautus, M., McLellan, S.L., Medema, G., Meschke, J.S., Mueller, J., Newton, R.J., Nilsson, D., Noble, R.T., Van Nuijs, A., Peccia, J., Perkins, T.A., Pickering, A.J., Rose, J., Sanchez, G., Smith, A., Stadler, L., Stauber, C., Thomas, K., Van Der Voorn, T., Wigginton, K., Zhu, K., Bibby, K., 2020. Wastewater-Based Epidemiology: Global Collaborative to Maximize Contributions in the Fight against COVID-19. Environ. Sci. Technol. 54, 7754–7757. https://doi.org/10.1021/acs.est.0c02388

Calderón-Franco, D., van Loosdrecht, M.C.M., Abeel, T., Weissbrodt, D.G., 2021. Free-floating extracellular DNA: Systematic profiling of mobile genetic elements and antibiotic resistance from wastewater. Water Res. 189. https://doi.org/10.1016/j.watres.2020.116592

Chik, A.H.S., Glier, M.B., Servos, M., Mangat, C.S., Pang, X.L., Qiu, Y., D’Aoust, P.M., Burnet, J.B., Delatolla, R., Dorner, S., Geng, Q., Giesy, J.P., McKay, R.M., Mulvey, M.R., Prystajecky, N., Srikanthan, N., Xie, Y., Conant, B., Hrudey, S.E., 2021. Comparison of approaches to quantify SARS-CoV-2 in wastewater using RT-qPCR: Results and implications from a collaborative inter-laboratory study in Canada. J. Environ. Sci. (China) 107, 218–229. https://doi.org/10.1016/j.jes.2021.01.029

Foladori, P., Cutrupi, F., Segata, N., Manara, S., Pinto, F., Malpei, F., Bruni, L., La Rosa, G., 2020. SARS-CoV-2 from faeces to wastewater treatment: What do we know? A review. Sci. Total Environ. https://doi.org/10.1016/j.scitotenv.2020.140444

Graham, K.E., Loeb, S.K., Wolfe, M.K., Catoe, D., Sinnott-Armstrong, N., Kim, S., Yamahara, K.M., Sassoubre, L.M., Mendoza Grijalva, L.M., Roldan-Hernandez, L., Langenfeld, K., Wigginton, K.R., Boehm, A.B., 2021. SARS-CoV-2 RNA in Wastewater Settled Solids Is Associated with COVID-19 Cases in a Large Urban Sewershed. Environ. Sci. Technol. 55, 488–498. https://doi.org/10.1021/acs.est.0c06191

Havelaar, A.H., Hogeboom, W.M., Pot, R., 1985. F specific RNA bacteriophages in sewage: Methodology and occurrence. Water Sci. Technol. 17, 645–655. https://doi.org/10.2166/wst.1985.0167

Hokajärvi, A.M., Rytkönen, A., Tiwari, A., Kauppinen, A., Oikarinen, S., Lehto, K.M., Kankaanpää, A., Gunnar, T., Al-Hello, H., Blomqvist, S., Miettinen, I.T., Savolainen-Kopra, C., Pitkänen, T., 2021. The detection and stability of the SARS-CoV-2 RNA biomarkers in wastewater influent in Helsinki, Finland. Sci. Total Environ. 770, 145274. https://doi.org/10.1016/j.scitotenv.2021.145274

Joonaki, E., Hassanpouryouzband, A., Heldt, C.L., Areo, O., 2020. Surface Chemistry Can Unlock Drivers of Surface Stability of SARS-CoV-2 in a Variety of Environmental Conditions. Chem. https://doi.org/10.1016/j.chempr.2020.08.001

Kattur Venkatachalam, A.R., Szyporta, M., Kiener, T.K., Balraj, P., Kwang, J., 2014. Concentration and purification of enterovirus 71 using a weak anion-exchange monolithic column. Virol. J. 11, 1–8. https://doi.org/10.1186/1743-422X-11-99

Kitajima, M., Ahmed, W., Bibby, K., Carducci, A., Gerba, C.P., Hamilton, K.A., Haramoto, E., Rose, J.B., 2020. SARS-CoV-2 in wastewater: State of the knowledge and research needs. Sci. Total Environ. https://doi.org/10.1016/j.scitotenv.2020.139076

Kramberger, P., Honour, R.C., Herman, R.E., Smrekar, F., Peterka, M., 2010. Purification of the Staphylococcus aureus bacteriophages VDX-10 on methacrylate monoliths. J. Virol. Methods 166, 60–64. https://doi.org/10.1016/j.jviromet.2010.02.020

Larsen, D.A., Wigginton, K.R., 2020. Tracking COVID-19 with wastewater. Nat. Biotechnol. 38, 1151–1153. https://doi.org/10.1038/s41587-020-0690-1

Lundy, L., Fatta-Kassinos, D., Slobodnik, J., Karaolia, P., Cirka, L., Kreuzinger, N., Castiglioni, S., Bijlsma, L., Dulio, V., Deviller, G., Lai, F.Y., Alygizakis, N., Barneo, M., Baz-Lomba, J.A., Béen, F., Cíchová, M., Conde-Pérez, K., Covaci, A., Donner, E., Ficek, A., Hassard, F., Hedström, A., Hernandez, F., Janská, V., Jellison, K., Hofman, J., Hill, K., Hong, P.-Y., Kasprzyk-Hordern, B., Kolarević, S., Krahulec, J., Lambropoulou, D., de Llanos, R., Mackulak, T., Martinez-García, L., Martínez, F., Medema, G., Micsinai, A., Myrmel, M., Nasser, M., Niederstätter, H., Nozal, L., Oberacher, H., Očenášková, V., Ogorzaly, L., Papadopoulos, D., Peinado, B., Pitkänen, T., Poza, M., Rumbo-Feal, S., Sánchez, M.B., Székely, A.J., Soltysova, A., Thomaidis, N.S., Vallejo, J., van Nuijs, A., Ware, V., Viklander, M., 2021. Making Waves: Collaboration in the time of SARS-CoV-2 - rapid development of an international co-operation and wastewater surveillance database to support public health decision-making. Water Res. 199, 117167. https://doi.org/10.1016/j.watres.2021.117167

Medema, G., Been, F., Ruijgers, H., KWR, 2020a. De SARS-CoV-2 rioolwatermonsters van 12 augustus. KWR [WWW Document]. Nieuws. URL https://www.kwrwater.nl/actueel/de-nieuwste-rioolwatermonsters/

Medema, G., Heijnen, L., Elsinga, G., Italiaander, R., Brouwer, A., 2020b. Presence of SARS-Coronavirus-2 RNA in Sewage and Correlation with Reported COVID-19 Prevalence in the Early Stage of the Epidemic in the Netherlands. Environ. Sci. Technol. Lett. 7, 511–516. https://doi.org/10.1021/acs.estlett.0c00357

Ooi, E.E., Low, J.G., 2020. Asymptomatic SARS-CoV-2 infection. Lancet Infect. Dis. https://doi.org/10.1016/S1473-3099(20)30460-6

Parra Guardado, A.L., Sweeney, C.L., Hayes, E., Trueman, B.F., Huang, Y., Jamieson, R.C., Rand, J.L., Gagnon, G.A., Stoddart, A.K., 2020. Development and optimization of a new method for direct extraction of SARS-CoV-2 RNA from municipal wastewater using magnetic beads. medRxiv. https://doi.org/10.1101/2020.12.04.20237230

Rabenau, H.F., Cinatl, J., Morgenstern, B., Bauer, G., Preiser, W., Doerr, H.W., 2005. Stability and inactivation of SARS coronavirus. Med. Microbiol. Immunol. 194, 1–6. https://doi.org/10.1007/s00430-004-0219-0

Randazzo, W., Cuevas-Ferrando, E., Sanjuán, R., Domingo-Calap, P., Sánchez, G., 2020a. Metropolitan wastewater analysis for COVID-19 epidemiological surveillance. Int. J. Hyg. Environ. Health. https://doi.org/10.1016/j.ijheh.2020.113621

Randazzo, W., Truchado, P., Cuevas-Ferrando, E., Simón, P., Allende, A., Sánchez, G., 2020b. SARS-CoV-2 RNA in wastewater anticipated COVID-19 occurrence in a low prevalence area. Water Res. 181. https://doi.org/10.1016/j.watres.2020.115942

Rimoldi, S.G., Stefani, F., Gigantiello, A., Polesello, S., Comandatore, F., Mileto, D., Maresca, M., Longobardi, C., Mancon, A., Romeri, F., Pagani, C., Cappelli, F., Roscioli, C., Moja, L., Gismondo, M.R., Salerno, F., 2020. Presence and infectivity of SARS-CoV-2 virus in wastewaters and rivers. Sci. Total Environ. 744. https://doi.org/10.1016/j.scitotenv.2020.140911

Rockey, N., Arts, P.J., Li, L., Harrison, K.R., Langenfeld, K., Fitzsimmons, W.J., Lauring, A.S., Love, N.G., Kaye, K.S., Raskin, L., Roberts, W.W., Hegarty, B., Wigginton, K.R., 2020. Humidity and deposition solution play a critical role in virus inactivation by heat treatment on N95 respirators. medRxiv 5, 1–14. https://doi.org/10.1101/2020.06.22.20137448

Schrader, C., Schielke, A., Ellerbroek, L., Johne, R., 2012. PCR inhibitors - occurrence, properties and removal. J. Appl. Microbiol. 113, 1014–1026. https://doi.org/10.1111/j.1365-2672.2012.05384.x

Tanhaei, M., Mohebbi, S.R., Hosseini, S.M., Rafieepoor, M., Kazemian, S., Ghaemi, A., Shamloei, S., Mirjalali, H., Asadzadeh Aghdaei, H., Zali, M.R., 2021. The first detection of SARS-CoV-2 RNA in the wastewater of Tehran, Iran. Environ. Sci. Pollut. Res. https://doi.org/10.1007/s11356-021-13393-9

Westhaus, S., Weber, F.A., Schiwy, S., Linnemann, V., Brinkmann, M., Widera, M., Greve, C., Janke, A., Hollert, H., Wintgens, T., Ciesek, S., 2021. Detection of SARS-CoV-2 in raw and treated wastewater in Germany – Suitability for COVID-19 surveillance and potential transmission risks. Sci. Total Environ. 751, 141750. https://doi.org/10.1016/j.scitotenv.2020.141750

Wu, F., Zhang, J., Xiao, A., Gu, X., Lee, W.L., Armas, F., Kauffman, K., Hanage, W., Matus, M., Ghaeli, N., Endo, N., Duvallet, C., Poyet, M., Moniz, K., Washburne, A.D., Erickson, T.B., Chai, P.R., Thompson, J., Alm, E.J., 2020. SARS-CoV-2 Titers in Wastewater Are Higher than Expected from Clinically Confirmed Cases. mSystems. https://doi.org/10.1128/msystems.00614-20

