## Supplementary Material for "Monitoring SARS-CoV-2 in sewage: toward sentinels with analytical accuracy"

Deposited as pre-print on *medRxiv*

### Supplementary material

#### Methods

##### qPCR analysis

We performed qPCR analysis, according to a previous study (Agrawal et al., 2021), using the TaqPath COVID-19 RT-PCR Kit (Thermo Fisher Scientific) which includes: (1) TaqPath COVID-19 Assay Multiplex, which contains three primer/probe sets specific to different SARS-CoV-2 genomic regions (i.e. N gene, S gene and Orf1ab gene) and primers/probes for bacteriophage MS2. (2) MS2 Phage Control – RNA control, having a concentration of 10^6 copies per µl, to verify the efficacy of the RNA extraction and the absence of inhibitors in the PCR reaction. (3) TaqPath COVID-19 Control – Positive SARS-CoV-2 RNA control that contains targets specific to the SARS-CoV-2 genomic regions targeted by the assays. The manufacturer (Thermo Fisher Scientific) has not publicly released the primers/probe sets sequences, therefore, we do not have access to the information related to the primers/probe sets sequences and the length of the PCR products.

**Table S1. Information about dyes corresponding to each target gene.**

| Gene | Dye | Quencher |
|---|---|---|
| ORF1ab | FAM | QSY |
| N Protein | VIC | QSY |
| S Protein | ABY | QSY |
| MS2 (Internal Control) | JUN | QSY |

In case of the positive control, we included triplicates of the four different concentration (i.e. 1x10^1, 2x10^1, 2x10^2, 2x10^3 copies per reaction) of the TaqPath COVID-19 positive control for each qPCR run. For MS2 phage internal control triplicates of the three different concentrations (i.e. 2x10^2, 2x10^3, 2x10^4 copies per reaction) were also included in each qPCR run. For the SARS-CoV-2 positive control and MS2 phage internal control, each reaction contained 12.5 µL TaqPath 1-Step Multiplex Master Mix (4X), 2.5 µL COVID-19 Real Time PCR Assay Multiplex, 33 µL nuclease free water, and 2 µL of positive or internal control. Triplicates of negative controls were also included in each run, each reaction contained 12.5 µL TaqPath 1-Step Multiplex Master Mix (4X), 2.5 µL COVID-19 Real Time PCR Assay Multiplex, and 35 µL nuclease free water. Ct values of positive control dilutions were plotted against known concentrations of the SARS-CoV-2 positive control and MS2 phage internal control, to generate standard curves. The start baseline value was set at 5 and threshold cycle (Ct) values were determined manually while adjusting the threshold to be above any background signal and within the exponential phase of the fluorescence curves. Primer efficiencies were 100.17 ± 5.98% for N, 103.71 ± 11.12% for S, 103.08 ± 5.42% for Orf1ab, and 86.42 ± 6.46% for MS2 phage (n = 3 runs, mean ± SD). The slopes of the standard curves for the quantification were −3.33 ± 0.14 for N, −3.22 ± 0.21 for S, −3.27 ± 0.14 for Orf1ab, and −3.67 ± 0.19 for MS2 phage. The SARS-CoV-2 concentrations detected in the samples are presented without correcting for recovery efficiencies.

**Table S2. PCR protocol and thermal profiles**

| Step | | Temperature | Duration |
|---|---|---|---|
| **Hold-Stage** | | 25 °C | 2 min |
| **Hold-Stage** | | 53 °C | 10 min |
| | | 95 °C | 2 min |
| **PCR-Stage** | **45 cycles** | 95 °C | 15 s |
| | | 60 °C | 1 min |

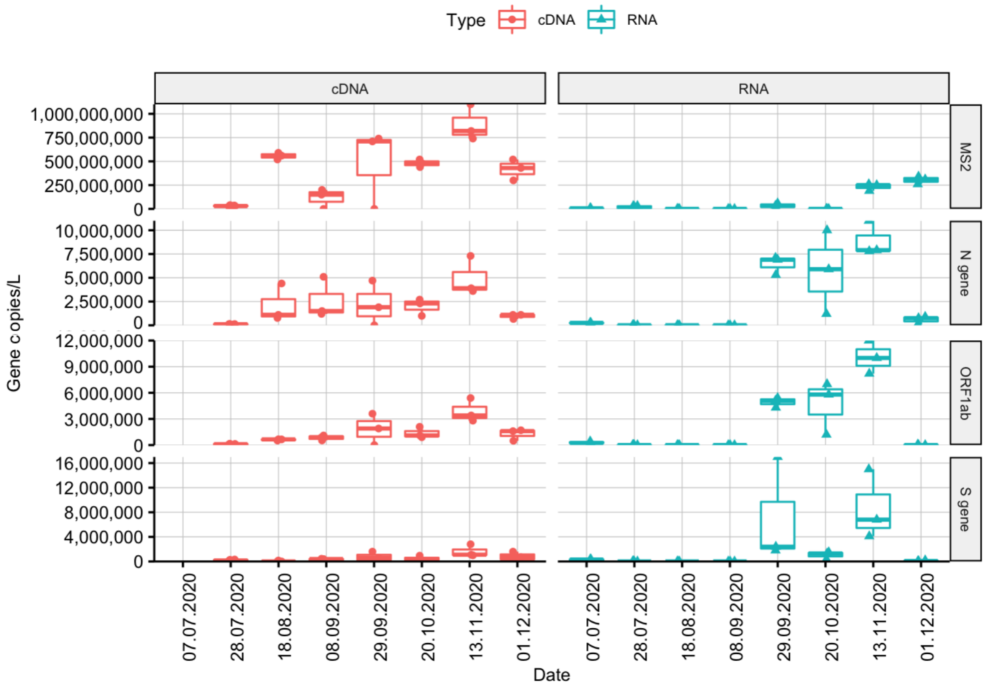


**Figure S1.** Concentration of SARS-CoV-2 RNA in influent wastewater as determined by the N, ORF1ab and S gene assays in gene copies per L using one-step RT-qPCR (RNA) or two-step RT-qPCR (cDNA) over the whole sampling campaign (July 2020 until December 2020) covering the second wave in The Netherlands.

#### Reference

**(1)** *TaqPath COVID-19 CE-IVD RT-PCR Kit Instruction for Use*; MAN0019215; Thermofisher Scientific.
